# Conducting a survey of abortion-related knowledge, attitudes and practices amongst health professionals in Britain, strategies adopted and lessons learned: evidence from the SACHA Study

**DOI:** 10.1101/2024.04.19.24306065

**Authors:** RS French, MJ Palmer, O McCarthy, N Salaria, R Meiksin, J Shawe, M Lewandowska, R Scott, K Wellings, the SACHA Study Team

**Affiliations:** Department of Public Health, Environments and Society, Faculty of Public Health and Policy, London School of Hygiene and Tropical Medicine, 15-17 Tavistock Place, London WC1H 9SH; School of Nursing and Midwifery, Faculty of Health, University of Plymouth

**Keywords:** survey, abortion, health professional, general practice, midwifery, sexual and reproductive health, pharmacy, Britain

## Abstract

**Background:** Most surveys examining health professionals’ knowledge, attitudes and practices around abortion have used convenience samples and have targeted doctors. Our goal in the SACHA Study, drawing on evidence-based strategies to maximise response rates, was to achieve a representative sample of a wider range of health professionals, working in general practice, maternity services, pharmacies, sexual and reproductive health (SRH) clinics and specialist abortion services in Britain, to explore the knowledge, attitudes and experience of abortion care and views on future models of delivery.

**Methods:** A cross-sectional questionnaire-based survey of midwives, doctors, nurses and pharmacists in England, Scotland and Wales was undertaken between November, 2021 and July, 2022. We used a stratified cluster sampling approach to select a random sample of sites and all eligible staff within those services were asked to respond to the survey. Evidence-based strategies to maximise completion rates were adopted, including postal delivery of the one-page questionnaire with personal letter of invitation and a stamped address envelope for return, inclusion of an unconditional voucher and follow-up.

**Results:** Overall, 147 of the 314 (46.8%) health service sites randomly selected took part in the survey. Reasons for non-participation included local Research and Development (R&D) Department non-response, lack of resources or insufficient time to support or approve the study, lack of interest in or perceived relevance of the topic and insufficient capacity to take part, exacerbated by work demands during the COVID epidemic. Of the 1370 questionnaires sent to eligible identified participants within these services, 771 were completed and returned (56.3%). At the service level the highest proportion of returns was from SRH clinics (81.0%) and the lowest from general practice (32.4%). In relation to profession, returns were highest amongst midwives (69.6%) and lowest amongst pharmacists (36.5%).

**Conclusions:** Obtaining information about health professional knowledge, attitudes and practices is key to guide service development and policy and to identify gaps in training and service provision in abortion. Despite our attempts to gain a representative sample of health professionals, the challenges we experienced limited the representativeness of the sample, despite the use of an evidence-based strategy.

## Background

More patients in Britain are having early medication abortions at home, facilitated by consultations conducted remotely via video or phone.^1,2^ These developments provide opportunities to examine which types of health services and professionals deliver, or could deliver with changes in regulations, abortion advice and care. In Britain, the 1967 Abortion Act only permits registered medical practitioners working in licensed abortion clinics or NHS hospitals to authorise abortions, prescribe abortion medication or perform abortion procedures, such as manual vacuum aspiration.^3^ Obtaining information on health professional knowledge, attitudes and practices is key to guide service development and policy and to identify gaps in training and service provision in abortion, or any health area. Failing to consult health service staff about potential changes to practice and policy is arguably unethical and may result in inefficiencies and, potentially, barriers to adoption of change. However, demanding work schedules, conflicting priorities, frequency of requests to take part in surveys and “gatekeepers” hindering access can reduce health professionals’ participation in reseach.^4^

Most health professional surveys on abortion care in Britain have focused on attitudes, particularly towards abortion law and conscientious objection.^5-13^ Other topics have included medical students’ future willingness to participate in abortion care,^5,6^ referral practices,^7^ views on models of service provision,^13,14^ and terminology.^15^ These surveys have been predominantly confined to doctors, including medical students,^9,10,12^ general practitioners (GPs),^7,9,10^ and obstetricians and gynaecologists.^9,11,12^ A few surveys of participants at specialist meetings or conferences have included doctors working in community sexual and reproductive health (SRH) clinics, nurses, and midwives, but overall representation of these professionals is scarce.^13-15^ The majority of surveys have used convenience samples, increasing the potential for bias. Recruitment from universities, conferences and general practices more actively involved in research is likely to reduce generalisability of findings to the wider workforce. Some studies have randomly selected a subset of those identified from professional organisations, including GPs via the British Medical Association,^10^ and obstetricians and gynaecologists via the Royal College of Obstetrics and Gynaecologists (RCOG).^11,12^ While these surveys, all distributed by post, have achieved high response rates (>70%), little information is provided about the process of random selection or on the socio-demographic characteristics of the respondents and the extent to which they were representative of the intended target group. Other surveys of health professionals designed to achieve a representative sample have achieved much lower response rates. For example, an online survey of GPs recruited via the Royal College of General Practitioners (RCGP) was reported to be broadly representative of members’ characteristics, but the estimated response rate was between 7-10%.^16^

Strategies to increase response rates have been shown to reduce bias. A Cochrane review on strategies used to increase survey response rates found the odds of response were at least doubled with monetary incentives, recorded postal delivery, a ‘teaser’ on the envelope to encourage respondents to open it, and an ‘interesting’ topic.^17^ To a lesser extent, odds of response were also significantly increased with pre-notification, follow-up, unconditional incentives, shorter questionnaires, sending out the questionnaire again at follow-up, mentioning an obligation to respond, university sponsorship, non-monetary incentives, personalised questionnaires, handwritten envelopes, inclusion of stamped address return envelopes, assurance of confidentiality and first class outward mailing. Systematic reviews of methods to improve survey response rates specifically amongst doctors and nurses have found similar findings,^4^ and also noted that endorsement from professional organisations increased response rates.^18,,19^ While postal and telephone surveys were more successful than online surveys, health professionals did respond well to having different options for questionnaire completion. Surveys of a sensitive nature have been found to have lower response rates.^4,17^ Amongst GPs, being too busy and lack of financial payment are reported as the most common reasons for non-response.^20^

The goal of the SACHA (Shaping Abortion for Change) Study was to provide an evidence base to inform optimal configuration of health services for the delivery of abortion care and provision in Britain. A component of this research was a survey of health professionals to assess their knowledge, attitudes and practices relating to abortion care. Our goal, drawing on evidence-based strategies to maximise response rates, was to achieve a representative sample of British health professionals working in general practice, maternity services, pharmacies, SRH clinics and specialist abortion services. In this paper we describe our approach, the extent to which the methods used were successful and what challenges were met.

## Methods

We conducted a cross-sectional questionnaire-based survey of health professionals working in primary and secondary health services in England, Scotland and Wales.

### Sampling and recruitment

Health professionals who have, or could potentially have, a role in providing abortion care and support were eligible to take part. These included: midwives, doctors, nurses and pharmacists currently working (either permanently or as a locum) in the following types of services: general practices, SRH services, pharmacies, maternity services, and abortion services (for maternity services, only midwives were eligible to take part). All eligible participants were required to be working in premises with postcodes and to be providing direct patient care, either face-to-face or remotely (via video-conferencing software or phone).

With the aim of achieving a representative sample, we used a stratified cluster sampling approach to identify services from which participants were to be recruited. A random sample of services, which constituted our ‘clusters’, was selected with all eligible staff within that service asked to respond. The only exception to this was midwives working in maternity services. Given larger numbers working across maternity services at each site, all midwives working within a 24-hour period identified by the site manager in either antenatal, labour and postnatal wards were eligible. To ensure adequate representation of health professionals in each of England, Scotland, and Wales, these three countries constituted our strata. Furthermore, to ensure proportional regional spread of services across England and to benefit from the precision gains associated with implicit stratified sampling,^23^ each sampling frame in England was ordered according to region (London, the North East, North West, Yorkshire, East Midlands, West Midlands, South East, East of England and the South West) and services were selected using systematic random sampling.

The samples of each service type were drawn independently from one another using sampling frames as shown in Table 1.

**Table 1.** Sampling frame and eligibility by service type.

| **Service type** | **Sampling frame and service identification** | **Eligible staff** | **Identification of staff and mailout period** |
| --- | --- | --- | --- |
| **General Practices** | List of General Practices compiled based on publicly available information from: Care Quality Commission, England; Health Inspectorate Wales and NHS Inform Scotland | General practitioners<br>Midwives<br>Pharmacists<br>Practice Nurses | Identified via practice website<br>8<sup>th</sup> November, 2021 – 17<sup>th</sup> March, 2022 |
| **Abortion Providers** | List of abortion providers compiled from: abortion statistics reports published Department of Health and Social Care (which includes details of all services in England and Wales that reported abortions to the Chief Medical Officer in the last year); lists of services provided by BPAS, NUPAS, and MSI Reproductive Choices; for Scotland, communication with those involved in abortion provision. | Doctors<br>Midwives<br>Nurses | Identified via service manager<br>11<sup>th</sup> January, 2022 – 27<sup>th</sup> July, 2022 |
| **Maternity Service** | Sampling frame was a list of all six-digit postcodes in England, Wales, and Scotland. Selected postcodes were entered into the ‘find a service’ function on the NHS website | Midwives | Identified via service manager<br>28<sup>th</sup> June, 2022 – 26<sup>th</sup> July, 2022 |
| **Pharmacies** | A list of registered pharmacies in England, Scotland and Wales available from the General Pharmaceutical Council | Pharmacists | Identified via pharmacy website or via phone<br>29<sup>th</sup> November, 2021 – 25<sup>th</sup> March, 2022 |
| **SRH Clinics** | Sampling frame was a list of all six-digit postcodes in England, Wales, and Scotland. Selected postcodes were entered into the ‘find a service’ function on the NHS website | Doctors<br>Midwives<br>Nurses | Identified via service manager<br>10<sup>th</sup> May, 2022 – 29<sup>th</sup> July, 2022 |

For NHS hospital-based abortion providers to be eligible, abortion services had to provide at least 100 abortions each year, of which ≥80% were classified as being carried out under ‘Ground C’ of the Abortion Act (i.e “the pregnancy has NOT exceeded its 24th week and that the continuance of the pregnancy would involve risk, greater than if the pregnancy were terminated, of injury to the physical or mental health of the pregnant woman”). For abortion services commissioned by the NHS and provided by the independent sector, which included British Pregnancy Advisory Service (BPAS), MSI Reproductive Choices, and the National Unplanned Pregnancy Advisory Service (NUPAS), up-to-date lists of clinics were sought from the relevant website, or directly from the service. We were unable to construct complete sampling frames for maternity and SRH services, and we therefore adopted a different approach to sampling. In these services our sampling frame consisted of a complete list of all six-digit postcodes in England, Wales, and Scotland. On randomly selecting a postcode, it was entered into the ‘find a service’ function on the NHS website to identify the nearest SRH clinic and maternity service (and its full postal address, contact details and website). General practices and pharmacies were identified from publicly available lists.

When selecting the sample of each service type, we also randomly selected several batches of ‘back-up samples’ using the same approach. This enabled us to approach additional randomly selected services in Batches B and C if a whole site identified in the original Batch A was found not to exist or was an ineligible service or declined to participate. For each service type, we initially sampled 45 services to approach, with the exception of pharmacies, where we sampled 100 services due to the likely number of eligible staff per site being lower.

On the basis of the population size of England, Scotland, and Wales (approximately 56 million, 5 million, and 3 million respectively), and assuming we would require approximately 45 of each type of service (except pharmacies), proportionate stratification would result in 39 clusters in England (0.875*45), 4 clusters in Scotland (0.078*45), and 2 clusters in Wales (0.047*45). Therefore, in order to ensure an adequate sample size in each nation, we over-sampled in Scotland and Wales so that at least six of every service type was located in each, with equivalent proportional oversampling for pharmacies, and reduced the number of sites in England accordingly to maintain feasibility.

Prior research conducted in the UK suggests that 46% of a random sample of GPs felt that the decision for an abortion should be the woman’s only, rather than the requirement of either one or, as is current practice, two doctors’ signatures.^10^ We needed 1200 completed surveys to give us precision of +/- 3% around this estimate and our aim was to achieve a minimum of 100 respondents in each practitioner group. No evidence from Britain was found on SRH doctor, nurse or pharmacist views to inform the sample size at the time of protocol development.

Eligible professionals working within each selected service were identified from website staff details/profiles (GP practices and pharmacies) and via contacting service managers (all services except GP practices). Where NHS staff names, professional category and contact information were shared by managers these were supplied, with staff permission, to the research team in a password-protected spreadsheet. When staff members declined to have their contact details passed to the research team, information on the total number of potential eligible participants working at that service was sought, in order to calculate the denominator for our response rate. We also worked with Clinical Research Network Local Clinical Specialty Research Leads to promote the research and support recruitment with local NHS Trusts. National professional organisations, including the British Society of Abortion Care Providers, the Faculty of Sexual and Reproductive Healthcare, the RCGP, the Royal College of Midwives, the Royal College of Nursing and the Royal Pharmaceutical Society, were informed about the survey to increase knowledge and profile of the research. For the identification of health professionals working in the NHS sites (i.e. maternity services, NHS abortion providers and SRH clinics), a Participant Identification Centre (PIC) Agreement was required with the local Research and Development (R&D) Department for each site. As names of health professionals working with general practices were in the public domain via practice websites it was not necessary to set up PIC agreements for these sites.

#### Data collection

A fully structured questionnaire was developed (see supplementary file). Classificatory data were collected on socio-demographic characteristics; attitudes towards abortion, including legal and regulatory frameworks and demedicalisation; experience of abortion care and support; views on integrating abortion provision into routine care; perceptions of implications for their roles and workload; self-assessment of competence and needs for professional training; and awareness, use and/or opinion of novel strategies or approaches, such as telemedicine. Likert scales were used to scale response options for attitudinal statements. To avoid bias stemming from a tendency to agree with attitudinal statements (acquiescence bias), statements were formulated representing differing views. For example, when eliciting views on the extension of roles in abortion, opposing statements expressed advantages for the health professional (e.g. increased job satisfaction) versus disadvantages (e.g. increased burden of workload). The questionnaire was piloted with representatives from each service type and amendments were made to improve clarity of response options and to include other options as appropriate, e.g. “don’t know”.

Each questionnaire pack included a personal letter of invitation, a Participant Information Sheet explaining the purpose of the study and giving assurance of confidentiality, the questionnaire, a tea bag (to encourage participants to complete the questionnaire over a tea-break), an unconditional £10 shopping voucher as a thank you for their time and a stamped addressed envelope for return of the questionnaire to the research team. Packs were posted to all identified individual professionals within each service at their workplace. A postal survey was chosen as it has been shown to yield higher response rates. ^10-12^ Each professional was provided with a unique ID number, which was pre-recorded on their paper questionnaire and used in follow up emails. The ID number indicated country, type of service, site and batch, so that response rates could be calculated. To maximise response rates, we used strategies others have found effective: i) including the unconditional incentive in the first mail out; ii) following up with two emails or (where providers are based on-site) phone calls at fortnightly intervals following the initial mail out for non-responders and iii) limiting the questionnaire to a maximum of two sides of A4. In the letter of invitation and in follow-up emails health professionals were given the option of completing the questionnaire online (using Online Surveys, www.onlinesurveys.ac.uk).^17^ The follow up emails were also a way of reaching people who may have been working from home over the COVID-19 pandemic. Completion and submission of the paper or online questionnaire implied consent.

Identifying information (names and contact details) were stored on a secure LSHTM server separately from the survey responses and were password-protected. Paper questionnaires were stored in a locked filing cabinet in a locked room (accessible only to the study team). Name and contact details were only used for research fieldwork purposes and will be destroyed at the end of the study.

#### Data analysis

Information on professional role, type of service and country were linked to questionnaire data using ID numbers. Data were entered into Online Surveys either by the research team for received paper questionnaires or direct by the participant. These were exported, and then analysed in Stata 17. At a service level, response rates were calculated by country and service type. Individual completion rates (i.e. the proportion of identified health professionals who returned a completed questionnaire) were calculated by country, service type, and professional group. Percentages (and frequencies) with 95% confidence intervals (CI) for participant socio-demographic characteristics were calculated accounting for clustering in the analysis. Cells with a count of ≤5 have been suppressed in tables.

## Results

Overall, 147 health service sites out of the 314 (46.8%) randomly selected took part in the health professional survey (see Table 2). The main reasons for site non-participation included R&D Department non-response, inability to support the study due to lack of resources or insufficient time to approve the study (maternity and SRH sites); lack of interest in the study among identified sites; a belief that that it was not relevant; reluctance to share names of staff or insufficient capacity to take part in research. In three sites (all abortion providers), approval and agreement to take part were established but no questionnaires were returned. We were unable to contact some pharmacies and SRH clinics by phone to invite eligible staff to take part, despite multiple attempts.

**Table 2.** Recruitment from randomly selected sites by country and type of service (n, %)*^

|  | **General Practice** | **Maternity** | **Abortion** | **SRH Clinic** | **Pharmacy** | **Total** |
| --- | --- | --- | --- | --- | --- | --- |
| England | 29/35<br>(82.9%) | 9/33<br>(27.3%) | 25/33<br>(75.8%) | 14/33<br>(42.4%) | 31/95<br>(32.6%) | 108/229<br>(47.2%) |
| Scotland | 5/6<br>(83.3%) | 1/6<br>(16.7%) | 7/10<br>(70%) | 4/6<br>(66.7%) | 6/16<br>(37.5%) | 23/44<br>(52.3%) |
| Wales | 5/7<br>(71.4%) | 2/6<br>(33.3%) | 2/6<br>(33.3%) | 1/6<br>(16.7%) | 6/16<br>(37.5%) | 16/41<br>(39.0%) |
| Total | 39/48<br>(81.3%) | 12/45<br>(26.7%) | 34/49<br>(69.4%) | 19/45<br>(42.2%) | 43/127<br>(33.9%) | 147/314<br>(46.8%) |

\* Includes sites where at least one respondent returned a questionnaire and for sites requiring R&D approval all permissions were obtained
^ Three Batch A general practices were replaced with sites from the Batch B general practice list and 34 Batch A pharmacies were replaced from the Batch B and C pharmacy lists. The reasons for replacement of the originally selected Batch A sites included inability to make contact (n=17), refusal to take part (n=11), short-term locums only (n=4), invalid telephone number (n=3) and site closures (n=2). We had insufficient time for attempts to replace non-participating Batch A maternity, abortion or SRH sites due to delays experienced setting up approvals.

Site participation, defined by the participation of a least one respondent from the site, was highest amongst general practices (81.3%) and lowest amongst maternity services (26.7%). It was highest in Scotland, 52.3% of sites identified, and lowest in Wales, 39.0%.

Figure 1 illustrates the geographical spread of participating sites and individual completion rates. Of the 1370 questionnaires sent out to identified participants within these services, 771 were completed and returned (56.3%). The proportion of returns was highest in Scotland (65.2%), followed by England (56.9%) and Wales (43.2%). Services with the highest proportion of returns were SRH clinics (81.0%), followed by specialist abortion providers (78.7%), maternity services (67.4%), pharmacies (39.7%), and lastly general practice (32.4%). In relation to profession, completed returns were highest amongst midwives (69.6%), followed by nurses (62.3%), doctors (45.1%) and lowest amongst pharmacists (36.5%). On average there were four respondents per general practice site, 17 per maternity site, eight per abortion service, six per SRH clinic and one per pharmacy. It was not possible in some sites to identify how many staff were eligible via the service managers, therefore we were unable to calculate an overall participation rate.

**Figure 1.**
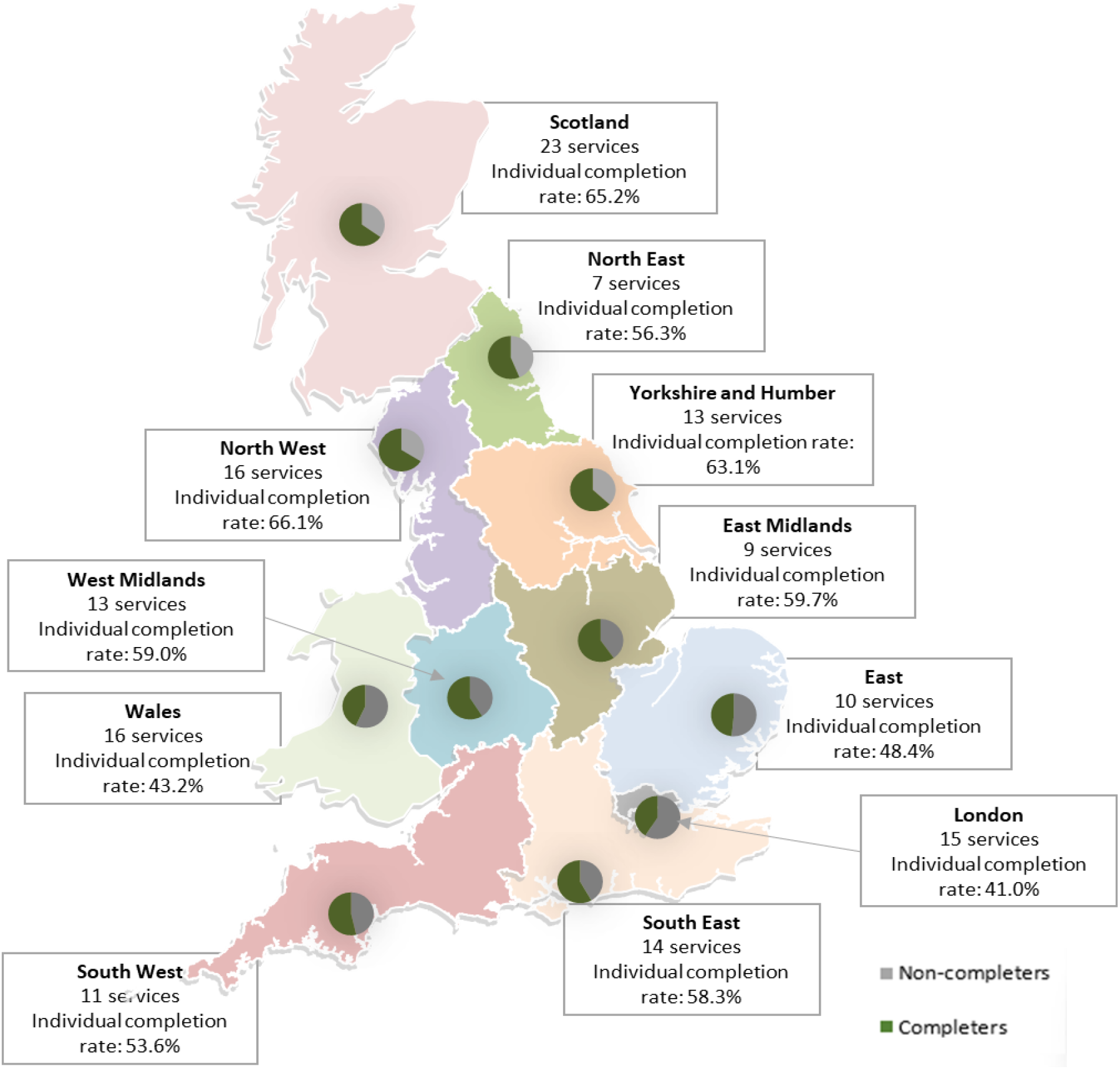
Site recruitment numbers and individual completion rates by region.

Examining the profile of participants by service illustrated some variations (see Table 3). The highest proportion of male health professionals was in pharmacies. Over half of those working in general practice and SRH services had been qualified for more than 20 years. Around a third of doctors and pharmacists reported right or centre leaning political beliefs. Over a half of those working in pharmacies and over a third of those working in general practice reported that religion was very or quite important in their lives.

**Table 3.** Characteristics of Survey participants by Service, n %, 95% CI.

|  | Total |  |  | General practice |  |  | Maternity |  |  | Abortion |  |  | SRH Services |  |  | Pharmacy |  |  |
| --- | --- | --- | --- | --- | --- | --- | --- | --- | --- | --- | --- | --- | --- | --- | --- | --- | --- | --- |
|  | n | % | 95% CI | n | % | 95% CI | n | % | 95% CI | n | % | 95% CI | n | % | 95% CI | n | % | 95% CI |
| **Total** | 771 |  |  | 156 | 20.26 | (13.36-29.51) | 198 | 25.71 | (13.32-43.80) | 247 | 32.08 | (21.19-45.34) | 115 | 14.94 | ( 8.22-25.61) | 54 | 7.01 | ( 4.66-10.42) |
| **Age** |  |  |  |  |  |  |  |  |  |  |  |  |  |  |  |  |  |  |
| Under 30 | 84 | 11 | ( 8.41-14.25) | † |  |  | 36 | 18.18 | (12.60-25.52) | 26 | 10.61 | ( 7.12-15.54) | 7 | 6.14 | ( 2.95-12.33) | 13 | 24.53 | (15.02-37.40) |
| 30-39 | 208 | 27.2 | (23.63-31.14) | 35 | 22.73 | (15.82-31.52) | 58 | 29.29 | (20.88-39.41) | 75 | 30.61 | (25.03-36.83) | 26 | 22.81 | (14.17-34.59) | 14 | 26.42 | (15.46-41.34) |
| 40-49 | 210 | 27.4 | (24.17-30.79) | 52 | 33.77 | (24.34-44.69) | 53 | 26.77 | (20.72-33.83) | 63 | 25.71 | (20.98-31.09) | 28 | 24.56 | (17.55-33.24) | 13 | 24.53 | (14.59-38.20) |
| 50 or over | 263 | 34.4 | (29.72-39.46) | 65 | 42.21 | (33.98-50.89) | 51 | 25.76 | (14.91-40.73) | 81 | 33.06 | (25.89-41.11) | 53 | 46.49 | (37.64-55.57) | 13 | 24.53 | (14.79-37.84) |
| **Gender** |  |  |  |  |  |  |  |  |  |  |  |  |  |  |  |  |  |  |
| Female | 669 | 87.6 | (83.52-90.70) | 112 | 72.73 | (65.66-78.81) | 197 | 100 |  | 222 | 90.98 | (85.74-94.42) | 105 | 92.11 | (86.13-95.64) | 32 | 60.38 | (45.99-73.17) |
| Male | 93 | 12.2 | ( 9.02-16.27) | 42 | 27.27 | (21.19-34.34) | † |  |  | 21 | 8.61 | ( 5.10-14.17) | 9 | 7.89 | ( 4.36-13.87) | 21 | 39.62 | (26.83-54.01) |
| Non-binary |  | † |  | † |  |  | † |  |  | † |  |  | † |  |  | † |  |  |
| **Professional role** |  |  |  |  |  |  |  |  |  |  |  |  |  |  |  |  |  |  |
| Doctor | 176 | 23 | (17.16-30.20) | 89 | 57.05 | (49.66-64.14) | † |  |  | 64 | 26.12 | (16.40-38.93) | 23 | 20 | (14.43-27.05) | † |  |  |
| Nurse | 261 | 34.2 | (25.67-43.81) | 58 | 37.18 | (29.86-45.13) | † |  |  | 112 | 45.71 | (32.57-59.48) | 89 | 77.39 | (68.85-84.13) | † |  |  |
| Midwife | 266 | 34.7 | (21.60-50.59) | 6 | 3.85 | ( 0.95-14.26) | 191 | 98.45 | (86.01-99.85) | 67 | 27.35 | (11.96-51.06) | † |  |  | † |  |  |
| Pharmacist | 62 | 8.12 | ( 5.54-11.73) | † |  |  | † |  |  | † |  |  | † |  |  | 54 | 100 | . |
| **When qualified** |  |  |  |  |  |  |  |  |  |  |  |  |  |  |  |  |  |  |
| Less than 5 years ago | 97 | 12.7 | ( 9.82-16.34) | 7 | 4.55 | ( 2.34- 8.65) | 34 | 17.26 | (10.29-27.49) | 36 | 14.75 | ( 9.76-21.69) | 9 | 7.89 | ( 4.47-13.57) | 11 | 20.75 | (11.13-35.39) |
| 5-10 years ago | 166 | 21.8 | (18.39-25.61) | 26 | 16.88 | (11.09-24.85) | 63 | 31.98 | (23.91-41.29) | 54 | 22.13 | (17.06-28.20) | 13 | 11.4 | ( 5.86-21.03) | 10 | 18.87 | (10.53-31.48) |
| 11-20 years ago | 216 | 28.2 | (25.12-31.53) | 39 | 25.32 | (19.23-32.57) | 48 | 24.37 | (18.06-32.01) | 79 | 32.38 | (27.12-38.12) | 34 | 29.82 | (21.19-40.18) | 15 | 28.3 | (16.94-43.31) |
| Over 20 years ago | 284 | 37.3 | (31.67-43.23) | 82 | 53.25 | (44.79-61.52) | 52 | 26.4 | (14.60-42.94) | 75 | 30.74 | (22.41-40.55) | 58 | 50.88 | (40.52-61.16) | 17 | 32.08 | (21.04-45.56) |
| **Country** |  |  |  |  |  |  |  |  |  |  |  |  |  |  |  |  |  |  |
| England | 560 | 72.7 | (59.64-82.79) | 119 | 76.28 | (57.17-88.57) | 172 | 86.87 | (50.74-97.70) | 148 | 59.92 | (35.62-80.16) | 84 | 73.04 | (30.61-94.33) | 37 | 68.52 | (51.09-81.93) |
| Wales | 79 | 10.3 | ( 4.67-21.05) | 16 | 10.26 | ( 3.49-26.53) | 25 | 12.63 | ( 2.10-49.33) | 7 | 2.83 | ( 0.45-15.74) | 24 | 20.87 | ( 2.73-71.25) | 7 | 12.96 | ( 5.47-27.70) |
| Scotland | 131 | 17 | ( 9.29-29.11) | 21 | 13.46 | ( 4.79-32.46) | † |  |  | 92 | 37.25 | (17.50-62.42) | 7 | 6.09 | ( 1.41-22.71) | 10 | 18.52 | ( 8.19-36.68) |
| **Importance of religion in life** |  |  |  |  |  |  |  |  |  |  |  |  |  |  |  |  |  |  |
| Very important | 60 | 7.87 | ( 6.07-10.15) | 18 | 11.61 | ( 7.20-18.20) | 13 | 6.67 | ( 3.63-11.93) | 13 | 5.31 | ( 3.06- 9.03) | † |  |  | 12 | 22.64 | (13.36-35.72) |
| Quite important | 147 | 19.3 | (16.51-22.42) | 38 | 24.52 | (19.20-30.75) | 40 | 20.51 | (14.16-28.76) | 34 | 13.88 | ( 9.77-19.34) | 20 | 17.54 | (11.58-25.69) | 15 | 28.3 | (17.76-41.91) |
| Not important | 520 | 68.1 | (63.81-72.12) | 92 | 59.35 | (50.61-67.54) | 138 | 70.77 | (59.16-80.18) | 185 | 75.51 | (68.35-81.49) | 82 | 71.93 | (59.97-81.43) | 22 | 41.51 | (27.33-57.25) |
| Prefer not to say | 36 | 4.72 | ( 3.45- 6.44) | 7 | 4.52 | ( 2.11- 9.42) | † |  |  | 13 | 5.31 | ( 3.05- 9.07) | 8 | 7.02 | ( 3.69-12.96) | † |  |  |
| **Political beliefs** |  |  |  |  |  |  |  |  |  |  |  |  |  |  |  |  |  |  |
| Right/right of centre | 30 | 3.95 | ( 2.71- 5.73) | 11 | 7.19 | ( 3.64-13.70) | † |  |  | 6 | 2.47 | ( 1.06- 5.66) | † |  |  | 6 | 11.32 | ( 5.18-22.99) |
| Centre | 114 | 14.9 | (12.50-17.64) | 36 | 23.53 | (18.53-29.40) | 21 | 10.71 | ( 5.94-18.57) | 29 | 11.93 | ( 8.59-16.34) | 16 | 14.04 | ( 9.21-20.81) | 11 | 20.75 | (12.12-33.22) |
| Left/left of centre | 227 | 29.9 | (26.38-33.70) | 48 | 31.37 | (24.32-39.40) | 64 | 32.65 | (26.20-39.84) | 78 | 32.1 | (24.54-40.73) | 27 | 23.68 | (15.42-34.57) | 10 | 18.87 | (10.37-31.85) |
| None | 303 | 39.9 | (35.71-44.29) | 39 | 25.49 | (18.87-33.47) | 93 | 47.45 | (41.87-53.10) | 95 | 39.09 | (33.02-45.52) | 59 | 51.75 | (38.19-65.07) | 17 | 32.08 | (21.07-45.51) |

† counts ≤5

A few participants left free text mentioning the value of the research and appreciating the opportunity to take part.

*This is a really important ongoing piece of work*. Midwife England

*I hope it* [the research] *improves care, support and choice for women!* Abortion service nurse England

The inclusion of the voucher and a tea bag in the questionnaire pack was also welcomed.

*Thank you for the teabag! I have passed on the £10 voucher to our staff who work tirelessly - much appreciated. :-)* GP Scotland

However, some practitioners returned their vouchers to the research team, with both completed and uncompleted questionnaires.

## Discussion

To date, this is the most comprehensive survey of health professionals’ abortion-related knowledge, attitudes and practices undertaken in Britain. Participants included nurses and midwives, whose views and experiences have been under-represented in prior surveys and, to our knowledge, this is the first survey about abortion amongst pharmacists in Britain. Previous British surveys of health professionals on abortion have, in the main, focused on medical students and doctors and used convenience samples or more localised populations to identify participants. Through random selection of service sites in Britain we aimed for a representative sample of eligible health professionals and through use of postal surveys, with an online completion option and an unconditional voucher we hoped to maximise the response rate.

Just under half of the sites identified through random sampling participated, with at least one eligible health professional responding. Site participation was lowest amongst NHS sites where we needed to set up PIC agreements before approaching staff, specifically maternity services and SRH clinics. Despite our requests from the participating sites, we were unable to obtain the numbers of eligible staff working in many of the SRH clinics, maternity services and abortion clinics, so were not able to obtain an overall denominator of eligible staff members for calculation of an overall participation rate. However, in terms of the completion rate amongst staff who were sent questionnaires, this was highest in SRH clinics and abortion services and lowest in general practice and pharmacies. Postal survey response rates amongst GPs are more commonly lower compared to specialist doctors and the evidence-base on strategies to increase GP response rates suggests they may have little effect.^22,23^ Amongst pharmacists, postal versus online surveys and the offer of vouchers versus not achieve higher response rates.^24,25^

Overall, responders to the survey were more likely to be female, qualified for more than 20 years and likely to report that religion was not important in their lives and they had no political beliefs. Workforce profiles show a higher proportion of females work across the health sector: 89% of nurses and midwifes,^26^ 62% of pharmacists,^27^ 60% of obstetricians and gynaecologists and 57% of GPs.^28^ The greater representation of females in our survey compared to males, may reflect the gender profile of staff working in the services that were included, but may be also due to females being more interested in the topic. The older age profile of our participants aligns with concerns that have been raised within primary care and SRH services about an aging workforce who are retiring and are not being replaced.^26,28,29^ With the exception of pharmacists, the majority of health professionals said that religion was not important in their lives. Reported religious affiliation and religiosity amongst medical students and nurses has been associated with more favourable attitudes towards conscientious objection.^5,8,30^ Around a quarter of GPs and pharmacists report having no religion.^27,28^

Despite our attempts to achieve a representative sample we experienced a number of challenges, which will affect the generalisability of our findings. First, many of the sites were experiencing severe staff shortages and huge workload demands. Fieldwork started in the general practice and pharmacy sites as PIC approvals were not required at these sites. However, questionnaires were sent out at a time when both of these services were responding to COVID. Fieldwork in the SRH clinics coincided with the mpox (monkeypox) outbreak. Second, local R&D approvals required in over 100 NHS sites was extremely time-consuming and resource intensive. Requirements for approval varied across departments and some sites were lost because of these delays or because the study was not viewed as a priority. As the end of the study approached there was insufficient time to replace these withdrawn sites with ‘Batch B’ ones. Third, identifying eligible health professionals in the selected sites was difficult. Staff turnover rates and reliance of locums were high in some services, and in general practice staff profiles on websites were sometimes out-of-date. Despite multiple attempts it was not possible to get through to some sites over the phone, particularly the pharmacies and SRH clinics. Some service managers were unwilling to provide the names of staff, even with their permission, despite reassurances that all ethical and R&D approvals were in place and that the dataset for analysis would be anonymised. Fourth, questionnaire packs were lost in the post and not received by sites, or questionnaires that staff said had been completed and posted back to the research team were never received. When individual work email addresses were available, online links to the survey were emailed to named staff. Sending out questionnaires by registered post may have helped, but many staff were still working at home or going into the workplace less frequently due to COVID. We did not always have individual staff email addresses for those working in general practices and pharmacies, and so had to rely on the phone for follow-up, in the case of general practice often via the practice manager. It was, therefore, in some cases not possible to confirm whether or not questionnaire packs had been received by those they had been posted to. This could have negatively impacted completion rates in these settings. Finally, despite explanations from the research team that we were seeking views of professionals not necessarily providing abortion advice or care, some managers explained that the topic was not relevant to their service or they did not agree with the topic so they would not be participating.

While it is essential that any research undertaken is ethical, unnecessary administrative procedures hinder research and result in a disproportionate amount of research funding being spent managing administration. For the SACHA health professional survey five members of the research team needed to be allocated to liaising with over 100 different R&D Departments to set up PIC agreements. The processes involved in gaining approvals for research involving the NHS seem to be based around the default assumption of any project being a randomised controlled trial involving patients. Our paper-based questionnaire to be completed by health professionals posed low risk to participants, however the regulatory frameworks we had to negotiate were no less cumbersome. A centralised system to obtain local approvals would have been much more efficient. We add our voice to calls have been made for greater harmonisation, simplifiication and proportionality of processes.^31^

A recent review concluded that even in the age of declining response rates, the accuracy of results based on random sample surveys is generally higher than that achieved from non-probability convenience samples.^32^ Even with the resource-intensive nature our approach, and the somewhat limited participation and completion rates, the findings reported from this sample are likely to be more reliable than had we simply opted for a convenience sample.

Despite our attempts to gain a representative sample of health practitioners, the challenges experienced reduced the effectiveness of the evidence-based strategy used. While the COVID pandemic undoubtedly affected the completion rates, the obstacles experienced in gaining locals approvals to survey health professionals and pressures within the NHS are likely to continue to affect future research. Given the crucial importance of professional opinion to improving health service provision, it is essential that ways are found to remove such obstacles and to facilitate the process of conducting empirical studies aimed at guiding health service reform.

## Data Availability

Survey data is not available as noted in Data Protection Impact Assessment. Study materials are available on request to the research team.

## List of abbreviations

BPAS: British Pregnancy Advisory Service
GPs: General Practitioners
NUPAS: National Unplanned Pregnancy Advisory Service
PIC: Participant Identification Centre
R&D: Research & Development
RCGP: Royal College of General Practitioners
RCOG: Royal College of Obstetricians and Gynaecologists
SACHA: Shaping Abortion for Change
SRH: Sexual & Reproductive Health

## Declarations

### Ethics approval

Approval for the survey was received from NHS Health Research Authority (IRAS Approval ID 297849), the LSHTM Ethics Committee (LSHTM Ethics Ref 26332), BPAS Research and Ethics Committee (reference number: 2021/08/FRE) and MSI Ethics Review Committee (application number 009-21).

### Author’s contributions

KW and RF conceptualised the study. RF, MP and JS designed the survey and materials, with input from all authors. RF, JS, MP, RM, and NS managed ethics/ R&D approvals and fieldwork. OM, RS and MP contributed to the analysis. RF drafted the paper with input from MP, RM, RS, NS, ML, JS and KW. All authors have read and approved the final manuscript.

## Acknowledgements

* Other members of the SACHA Study team include: Annette Aronsson (Karolinska Institute, Sweden), Paula Baraitser (Kings College London), Sharon Cameron (University of Edinburgh), Caroline Free (LSHTM), Louise Keogh (University of Melbourne, Australia), Patricia A. Lohr (BPAS), Clare Murphy (BPAS), Wendy V. Norman (University of British Columbia, Canada), Jennifer Reiter (Lambeth Local Authority), Sally Sheldon (University of Bristol) and Geoff Wong (University of Oxford).

We are grateful to all the survey participants who generously dedicated their time and shared their experiences, and to all service managers who helped identify eligible staff. We would like to thank the North Thames Clinical Research Network for their support and local R&D Departments who helped obtain approvals.

We also thank our Advisory Group for supporting the development of this study: Jonathan Lord (Chair), Marge Berer, Sue Mann, Claire Anderson, Joanne Fletcher, Becky Gunn, Charlotte Kelly, Tracey Masters, and Sam Rowlands.

